# Prognostic Stratification of Familial Hypercholesterolemia Patients Using AI Algorithms: A Gender-Specific Approach

**DOI:** 10.1101/2024.10.11.24315359

**Authors:** A Zamora, L Masana, F Civeira, D Ibarretxe, M Fanlo-Maresma, A Vila, Manuel Suárez Tembra, Marco-Benedí Victoria, Luis A. Alvarez-Sala-Walther, C Camacho, the National Registry of the Spanish Society of Arteriosclerosis

## Abstract

**Background:** Familial Hypercholesterolemia (FH) is the most prevalent autosomal dominant disorder, affecting about 1 in 200-250 individuals, with an estimated 30 million patients globally. It is the leading cause of early and aggressive coronary artery disease (CAD).

**Objective:** To developed an artificial intelligence (AI) and machine learning (ML) algorithm for cardiovascular risk stratification in a FH population, emphasizing sex-specific differences and model explainability.

**Methods:** We analyzed patients with genetically confirmed FH or a score greater than 8 on the Dutch Lipid Clinics Network (DLCN) criteria from the National Registry of the Spanish Atherosclerosis Society, including individuals enrolled from January 2010 to December 2017. The model utilized a comprehensive dataset incorporating family history, clinical characteristics, laboratory results, genetic data, imaging studies, and lipid-lowering treatment details. Eighty percent of the population was allocated for training the AI algorithm, while the remaining 20% was used for testing and 70/30 population for internal validation. A Histogram-based Gradient Boosting Classification Tree was used. The stability of the AI system was assessed using K-fold cross-validation. Shap methodology analyzed the influence of different variables by sex. Youden’s J statistic established the optimal cutoff point for identifying very high cardiovascular risk.

**Results:** A total of 1.764 patients were included (51.8% women), among whom 264 experienced major adverse cardiovascular events (MACE), with 8% being women. Notably, 52% experienced a cardiovascular event before age 50, and 37% had subclinical atherosclerosis. The final model incorporated 82 variables, achieving metrics of precision for MACE accuracy (0.92), recall (0.89), F1 score (0.91), and ROC (0.88; 95% CI, 0.85-0.90), revealing significant sex-based differences. Women showed a lower association with MACE compared to men, although this effect diminished with the inclusion of multiple variables, particularly in younger women. In the model, age, GGT levels, and subclinical disease significantly impacted risk for women, while year of birth, age at initiation of statin treatment and HbA1c levels were more influential for men. The optimal risk threshold was 0.25 for the association with of MACE

**Conclusion:** AI-ML algorithms are promising tools for enhancing vascular risk stratification in patients with FH, revealing critical sex-based differences.

## Introduction

Familial Hypercholesterolemia (FH) is a genetic disorder resulting in very high LDL-C and increased risk for CVD. It is the most common autosomal dominant disease, with a prevalence of around 1/200-250 in our environment. Currently, it is estimated that there are around 30 million patients suffering from FH worldwide. In addition, around 20% to 25% of the diagnosed individuals are children and teenagers. It represents the most common genetic cause of early and aggressive coronary artery disease (CAD). Without treatment, 50% of men under 50 and 30% of women under 60 with FH will primarily develop CAD. The life expectancy of individuals with FH has been calculated to be between 10 and 30 years lower compared to the non-FH population (1). FH patients have a 45% higher coronary mortality after myocardial infarction and their risk of recurrence is 2.5 times higher than the general population. Therefore, early identification is very important to optimize treatment and to reduce extreme cardiovascular risk. Unfortunately, FH is frequently underdiagnosed and often undertreated. Underdiagnosis and undertreatment of FH are partially due to the lack of an effective gold standard to identify high-risk patients at an early stage. Recently, both the World Heart Federation and the International FH Foundation have alerted that FH is a public health priority that requires global action to improve its diagnosis,its treatment and to reduce its impact on CVD. They recommend the development of new screening systems and the use of digital tools to improve patient risk stratification (2).

Risk assessment represents the first critical step in the current approach to primary prevention of atherosclerotic cardiovascular disease (ASCVD). Risk calculators cannot be used interchangeably as they have been shown to over- or underestimate cardiovascular risk in populations other than those from which they are derived (3).The cardiovascular risk scales commonly used in clinical practice, such as Systematic COronary Risk Evaluation Score (SCORE) (4), SCORE2 (5), and SCORE2-OP (6) underestimate the risk in FH population. In these scales, FH patients with additional cardiovascular risk factors or vascular disease are classified as very high risk. Those without these criteria are classified as high vascular risk.Currently, there are only three specific risk calculators for FH: Montreal-FH score designed in a Canadian population (7,8), SAFEHEART-RE (9), and SIDIAP-FHP (10). The first two are based on a genetically defined population with FH and SIDIAP-FH in patients with phenotypic FH.

While men and women share many traditional risk factors for CVD, additional gender-specific risk factors and mechanisms are at play. Therefore, it is crucial to consider gender differences when it comes to predicting and managing CVD risks. Men and women in general (11) and population with FH (12) differed in the impact of the individual risk factors on the development of ASCVD. Different studies have shown that women with FH are undertreated compared to men with the same risk ASCVD, even in secondary prevention probably due to the underestimation of cardiovascular risk in women (13, 14). None of the three FH-specific risk calculators are developed specifically from a sex–gender perspective (7–10).

Machine learning (ML) and artificial intelligence (AI) offer promising alternatives by integrating complex datasets and providing more personalized risk assessments. Recently, machine learning (ML) models have been widely used to precisely predict CVD risk factors and providing a new instrument to improve early identification high risk patient, determine a patient’s CVD prognosis, make better decisions in clinical practice and determinate a personalized treatment strategy (15).In the field of FH, the use of ML and artificial intelligence (AI) techniques is seen as a significant advancement for improving screening, diagnosis, and risk assessment based on various data sources, such as electronic health records, plasma lipid profiles, genetic studies, radiology images and corneal arcus images (16). Threfore, it is necessary to develop robust, explainable, reliable, and ethical AI algorithms. It is crucial that these new tools incorporate the sex-gender perspective and social determinants to avoid potential biases and to prevent the continuation of a lack of information on male-female differences in the new era of digital medicine (17).

The aim of this study is to develop a ML-AI algorithm from a sex-gender perspective useful for cardiovascular risk stratification in FH population and with a significant emphasis on model of explainableartificial Intellingence (XAI) to provide maximum confidence to professionals and users. For the development of the model, data from family history, clinical, analytical, genetic, imaging data and, age at initiation of statin treatment, duration and intensity of lipid-lowering treatment from the Spanish Society of Arteriosclerosis registry were included.

## Methods

### Study design and population

In the present study, patients with diagnosis of FH, either with a positive genetic study or a Dutch Lipid Clinic Network score (DLCN) equal to or greater than 8, have been selected from the National Registry of the Spanish Society of Arteriosclerosis (SEA). These patients were included in the registry between January 2010 and December 2017.

The National Registry of Dyslipemias of the SEA is an online, retrospective, and prospective database (http://www.rihad.es) where accredited Spanish lipid units, recognized by the SEA, enter data from patients with lipid metabolism disorders. Established in 2013, the registry collected clinical, analytical, genetic, and follow-up data from 4.449 patients by 2017. The registry records a large volume of information in real-time, including sociodemographic data, family and personal medical history, analytical results with and without treatment, genetic data, and information on lipid-lowering and other treatments. This data, gathered under strict quality criteria from 60 lipid units across all 18 regions of Spain, is entered by qualified clinical professionals.

### Variables

The following baseline variables were obtained from the RIHAD database:

a) Personal and first-degree family history
Sex, paternal hypercholesterolemia, maternal hipercolesterole, family history of cardiovascular disease, age at first cardiovascular event in relatives, cardiopatíaisquémica, Ischemic heart disease, myocardial infarction, acute coronary síndrome, stable angina, coronary bypass, angioplasty, stroke, çstroke type, peripheral artery disease, aortic/abdominal aneurysm, Other cardiovascular events, aortic stenosis, hypertension, age at hypertension diagnosis, diabetes, smoker, Non-smoker,former smoker, Hepatic steatosis, packs/day per years of smoking.
b) Physical examination
BMI (Body Mass Index), waist, pulse, systolic blood pressure, diastolic blood pressure, tendinous xantoma, corneal arcus
c) Laboratory data: first visit to the lipid unit (baseline values without treatment) and current (the most recent available lab results with treatment)
Glucose, Uric acid, Lipoprotein (a), Gamma-glutamyl transferase; Alanine aminotransferase; Thyroid-stimulating hormone; Apolipoprotein A1; Apolipoprotein B; Glycated hemoglobin, Current total, Current HDL cholesterol; Current non-HDL cholesterol; Current LDL cholesterol; Current direct LDL cholesterol; Current triglycerides; Current glucose; Current creatinine; Current glomerular filtration rate; Current uric acid; Current GGT; Current ALT; Current TSH; Current bilirubin; Current glycated hemoglobin; Current microalbuminuria; Insulin, Homeostasis Model Assessment
d) Known subclinical arteriosclerosis: atherosclerotic lesions, incidentally detected by carotid echocardiography in asymptomatic persons
e) Total DLCN score (Dutch Lipid Clinic Network)
f) Genetic data
g) Data on treatment and follow-up in the Lipid Unit.
Age at statin initiation, lipid-lowering treatment, treatment duration, statin dose, total years on statins, total years on Ezetimibe, age at first visit at Lipid Unit

Definition of Major Adverse Cardiac Events (MACE).Presence of at least one of the following diagnoses:CVD, CAD, Ischemic heart disease, Myocardial infarction, Acute coronary síndrome, Stable angina, Coronary bypass, Angioplasty, Stroke, Stroke type, Peripheral artery disease, Aortic abdominal aneurysm, cardiovascular events.

The clinical diagnoses were extracted using the codes according to the International Classification of Diseases (ICD-10) from hospital discharge reports.

### Machine learning–based approach

We followed a model based on Cross-Industry Standard Process for Data Mining (CRISP-DM) (20). The model has 6 steps: problem understanding, data understanding, data preparation, modeling, quality assurance and explicability.

All AI-Machine Learning analyses and statistical analyses were conducted using Python programming language version 3.10.15 and its standard library.

Problem Understanding. The first step is to obtain a model that can assist to predict the risk of a MACE in a patient with FH. Our system must be able to take a information about the patient and return a MACE risk metric that ultimately classifies the patient in either Low Risk or High Risk. Data Understanding is divided into four tasks: requirement intake, data acquisition, data exploration and quality assurance. Data Preparation: cleaning, integration, feature engineering and scale. In our case, we discarded the following variables: Physical exercise frequency due to significant missing data. We apply the Standard Scale from sklearn library. This scaler applies the following transformation:

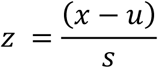

Where x is our sample, u is the mean of the training samples and s is the standard deviation of the training samples.

Modelling. We proposed a model for this problem: Histogram-based Gradient Boosting Classification Tree (HGBCT). A HGBCT is a machine learning algorithm used for classification tasks. It combines the principles of decision trees and boosting with the efficiency gained from binning continuous features into histograms, thus significantly improving performance on large datasets.The algorithm uses an ensemble of decision trees, where each tree corrects the errors of the previous ones. Boosting works by sequentially adding models (trees), with each new model focused on correcting the errors made by the previous ones. This helps in reducing bias and improving the model’s prediction accuracy (21). One of the main reasons to use it is that the estimator has native support for missing values.Once we had our model we configured it with the class_weight ‘balanced’ parameter. This means that the model uses the values of the target variable to automatically adjust weights inversely proportional to class frequencies in the input data. Data is then split into 5 folders following the Cross Validation principle with 20/80 test/train set to train the model with a Grid search for hyper parameter optimization. On top of this we make a scorer based on F1 score and pass it to the model. This way we optimize F1 score instead of Accuracy leading to better models when unbalanced classes are present. Once we have our model configured we train the model with 70% of data and save the remaining 30% for evaluation purposes.We ran a K-fold Cross Evaluation (22) with the base model to study the stability of the model for this problem. It helps ensure that the model generalizes well to unseen data by using different portions of the dataset for training and testing in multiple iterations. K-fold cross-validation was employed to ensure model stability by partitioning the dataset into k subsets, training on k-1, and testing on the remaining set (22). We ran the model 5 times with different combinations of data in the train and test set.

The evaluation metrics used for assessing the performance of the ML models included:

- Accuracy: This metric represents the proportion of correct predictions (both true positives and true negatives) out of the total number of predictions made by the model. It provides a general sense of the model’s performance but can be misleading in imbalanced datasets, where one class is significantly more prevalent than the other.
- Precision: Also known as positive predictive value, precision is the proportion of true positives out of all predicted positives (i.e., the fraction of correct positive predictions). High precision indicates a low false positive rate, meaning the model is good at avoiding incorrect positive predictions.
- Recall: This metric reflects the ability of the model to identify true positives, defined as the proportion of actual positives that the model correctly identifies. High recall indicates that the model is capturing a large percentage of the actual positive cases, though it may also produce false positives.
- F1-Score: It is the harmonic mean of precision and recall, providing a balanced measure when there is an uneven class distribution. It is particularly useful when both precision and recall are important, as it balances the trade-off between these two metrics.

These metrics provide a comprehensive evaluation of the machine learning models’ performance, particularly in the context of MACE in FH patients, where both false positives and false negatives must be carefully managed.

### Complementary analysisof Explicability by SHapley Additive exPlanations (SHAP)

It is important to explain how the model reaches a solution and what is the relationship of the different features with the output variable. One of the best algorithms to study explainability is SHAP. Shap analyze the contribution of each feature to the target variable per sample. In other words, we can estimate how any piece of data in our dataset affected the output given by the model. One way to represent this information is by plotting all samples per feature and see how they affect the outcome of the model (23,24). The SHAP summary plots demonstrate the either positive or negative adjustment to MACE risk estimation (x-axis) for each of the predictor variables (y-axis). Relative values for individual predictors were represented on a continuous color bar.

### Complementary analysis of Youden’s Jand ratio

The Youden’s *J*and ratio, the sum of sensitivity and specificity minus one, was used to establish the optimal cutoff point for the model associated with very high cardiovascular risk, considering that the cost is the same for a false positive as for a false negative. When using this index, one implicitly uses decision theory with a ratio of misclassification costs which is equal to one minus the prevalence proportion of the disease (25).

### Ethical aspects

The present study was approved by the institutional review board (Ethics and Medical Research Committee of Mataró Hospital, part of the Maresme Health Consortium, Mataró, Barcelona, Spain; Code 27/24).

## Results

### Study population

Of the 4.495 subjects in the study database at the time of inclusion, 2.685 patients were excluded due to diagnoses of combined hyperlipidemia, mixed hyperlipidemia, polygenic hyperlipidemia, unspecified dyslipidemia, or familial hypercholesterolemia without a positive genetic study or a DLCN score <8. Of the 1.764 subjects finally included, 1.540 had a positive genetic study. Among these, 95.6% had mutations in the LDLR gene, with 92.5% being simple heterozygotes, 4.1% compound heterozygotes, 2.1% double heterozygotes, and 1.2% homozygotes. The remaining subjects had a DLCN score ≥ 8.

Of the included patients, 127 (7.20%) were not Spain. Of the total included subjects, 838 (51.8%) were women. The mean age was 50 (±26) and 48 (± 15), in women and men respectively (p<0.02). A total of 264 living patients (73 women and 191 men) had a history of MACE (p<0.001). Among the patients, 36.2% had a family history of premature cardiovascular disease in first-degree relatives without sex differences. Additionally, 406 patients (37%) presented with subclinical atherosclerotic disease detected by imaging techniques. Of these, 190 were women and 216 were men (p<0.002).

Seventeen percent of the subjects had a history of hypertension, and 5.8% had diabetes, with no significant differences between sexes. Twenty percent of women and 23.8% of men were smokers (p<0,001). Men had a significantly higher body mass index (BMI) compared to women (p<0,001). Tendinous xanthomas were present in 39% of subjects, with no differences between sexes. The corneal arc was present in 29% of women and 39% of men (p < 0.001).The DLCN score was significantly lower in women (16 ± 4 vs. 17 ± 4, p < 0.001). The baseline LDL-cholesterol was 277 (±79.4) mg/dL with no differences by sex, while the post-treatment LDL cholesterol was 146 (±56.5) mg/dLin women and 139 (±61,9) mg/dL in men, respectively (p > 0.001).The atherogenic indices Apoprotein B/Apoprotein A1 and Triglicerides/HDL-Cholesterol were significantly higher in men (p < 0,001). The mean levels of Lipoproteina(a) were 29 (11–63) mg/dL, with no significant differences between sexes.

The mean age at the first visit to the Lipid Unit was 42 years (±15), with women being older than men (43 ± 16 vs. 42 ± 15 years) (p<0.02). Women were treated with lower doses of statins (p<0,001) and were less frequently prescribed combination therapy (statin + ezetimibe) (p<0.03). Additionally, lipid-lowering treatment was initiated later in women compared to men (p<0.006).

All data presented in this section are available in Table S1 (Supplementary materials).

### Machine learning models

The model included clinical, analytical, genetic and imaging variables, as well as age at treatment initiation, intensity, and use of combination therapy (statins + ezetimibe), totaling 82 variables.The variables included in the final algorithmic model are shown in Supplementary Table 2.

Figure 1 shows the Heatmap illustrating the relationship between variables and diferent cardiovascular disease or MACE in a population with FH.

**Fig. 1.**
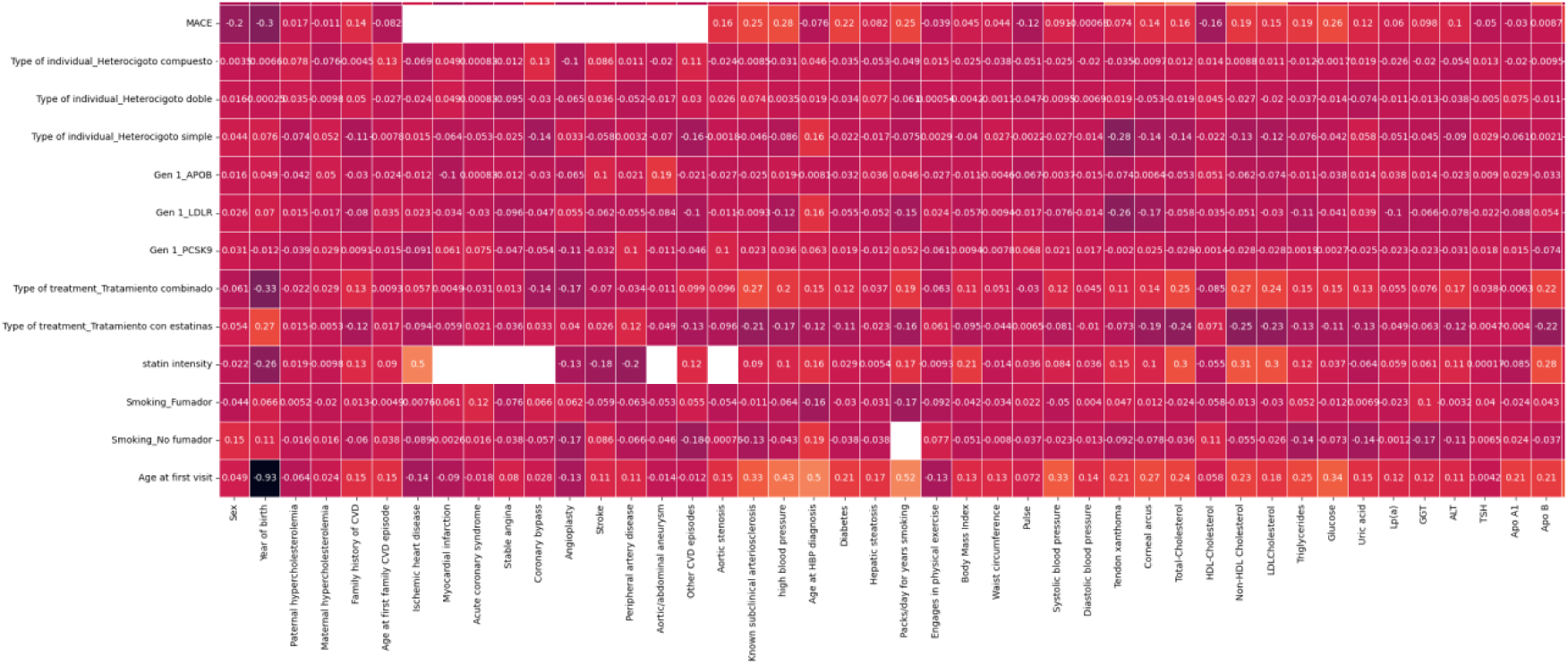
Heatmap illustrating the relationship between variables and diferent cardiovascular disease or MACE in a population with FH. The heatmap displays the strength of associations between selected variables— such as lipid levels, genetic markers, and treatment history—and the occurrence of MACE. Darker colors indicate stronger correlations, either positive or negative, while lighter colors represent weaker or no associations. A threshold for statistical significance was set at p < 0.05. This figure provides an overview of the most impactful factors contributing to cardiovascular risk in FH patients, facilitating targeted preventive strategies and personalized management.

Figure 2 presents the confusion matrices for the various machine learning models used to predict MACE in patients with FH. Each confusion matrix illustrates the performance of a distinct model, indicating true positives, true negatives, false positives, and false negatives in the prediction of MACE. The comparison includes results from 5-fold cross-validation. These matrices offer a visual comparison of each model’s predictive accuracy and misclassification rates, providing valuable insight into the selection of the optimal model for MACE prediction in FH patients.

**Fig 2.**
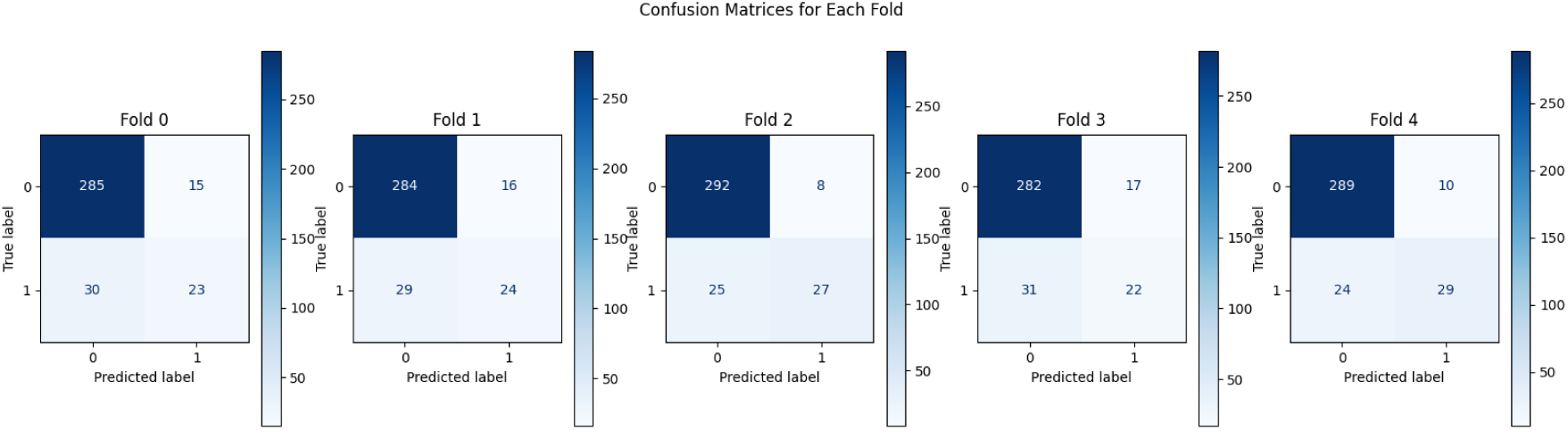
Confusion matrices for various machine learning models used in training to predict MACE in FH patients. Each confusion matrix displays the performance of a different machine learning model, highlighting true positives (TP), true negatives (TN), false positives (FP), and false negatives (FN) in the prediction of MACE. The models compared include 5 folds. These matrices provide a visual comparison of each model’s predictive power and potential misclassification rates, guiding the selection of the optimal model for MACE prediction in FH patients.

The various metrics obtained from the algorithmic model for the general population and when divided by sex are presented in Table 1. Notable differences in performance metrics are observed when the algorithm, trained on the general population, is applied separately to female and male cohorts. The AI algorithm model trained for the prediction of MACE demonstrated a recall of 0.98 in the female subpopulation and 0.82 in the male subpopulation for the presence of MACE. For the absence of MACE, the recall was 0.23 for women and 0.61 for men. The F1-score for MACE was 0.96 for women and 0.85 for men, while for the absence of MACE, the F1-scores were 0.32 and 0.55 for women and men, respectively. These results highlight significant differences in model performance between sexes, particularly in predicting the absence of MACE. Definition of the optimal cutoff point for the association with a pattern of high or low cardiovascular risk in FH population was assessed using the Youden Index.

**Table 1.** Performance metrics of the machine learning algorithm for predicting MACE and No MACE in patients with FH, presented globally and stratified by sex.

| Metrics | General | Women | Men |
| --- | --- | --- | --- |
| Accuracy MACE | 0,88 | 0,92 | 0,78 |
| Accuracy NO MACE | 0,84 | 0,92 | 0,78 |
| Precision MACE | 0,93 | 0,94 | 0,88 |
| Precision NO MACE | 0,47 | 0,56 | 0,50 |
| Recall MACE | 0,87 | 0,98 | 0,82 |
| Recall NO MACE | 0,65 | 0,23 | 0,61 |
| F1-Score MACE | 0,90 | 0,96 | 0,85 |
| F1-Score NO MACE | 0,54 | 0,32 | 0,55 |

\*The table reports key performance metrics, including accuracy, precision, recall, specificity and F1 score for the predictive model of MACE. Results are provided for the overall population and separately for male and female patients. These metrics enable comparison of the algorithm's performance across sexes, highlighting potential differences in predictive accuracy and model robustness in stratified subgroups. This detailed breakdown aids in understanding the generalizability and effectiveness of the model in diverse patient populations.

To define the optimal risk threshold, the population dataset used for model training was applied. Various thresholds were tested, assigning different weights to the error based on the Youden Index. The optimal risk threshold, where equal weight was given to both false positive and false negative errors, was set at 0.25 points to define very high or extreme risk for association with MACE (Fig. 3). The ROC curve for the AI algorithm, based on the cutoff point defined as optimal, is presented in Figure 4. This curve illustrates the algorithm’s performance at the selected threshold, highlighting the balance between sensitivity and specificity in predicting the occurrence of MACE.

**Fig. 3.**
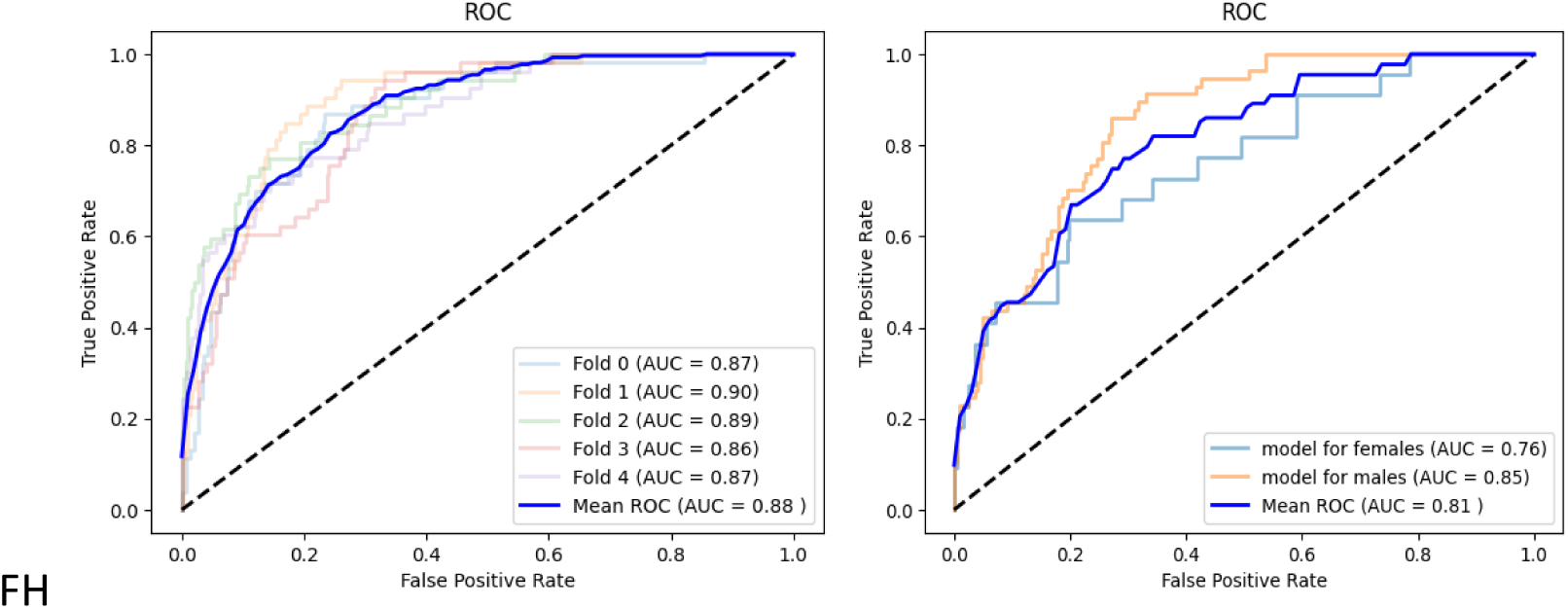
Receiver Operating Characteristic (ROC) curve using the K-fold cross-validation method to assess the robustness of the model predicting MACE in a population with FH. a) The ROC curve illustrates the performance of the predictive model across multiple K-folds, with the Area Under the Curve (AUC) representing the model’s ability to distinguish between patients who experienced MACE and those who did not. Each fold represents a different subset of the data used for training and validation, ensuring the model’s generalizability and robustness. The model demonstrates consistent AUC values across folds, supporting its reliability in predicting cardiovascular risk in FH patients. b) The ROC curves compare the model’s performance in predicting MACE separately for male and female patients. The Area Under the Curve (AUC) is shown for each sex, indicating the model’s discriminative ability in both populations. Differences in AUC values highlight potential sex-specific variations in risk prediction accuracy

**Fig. 4.**
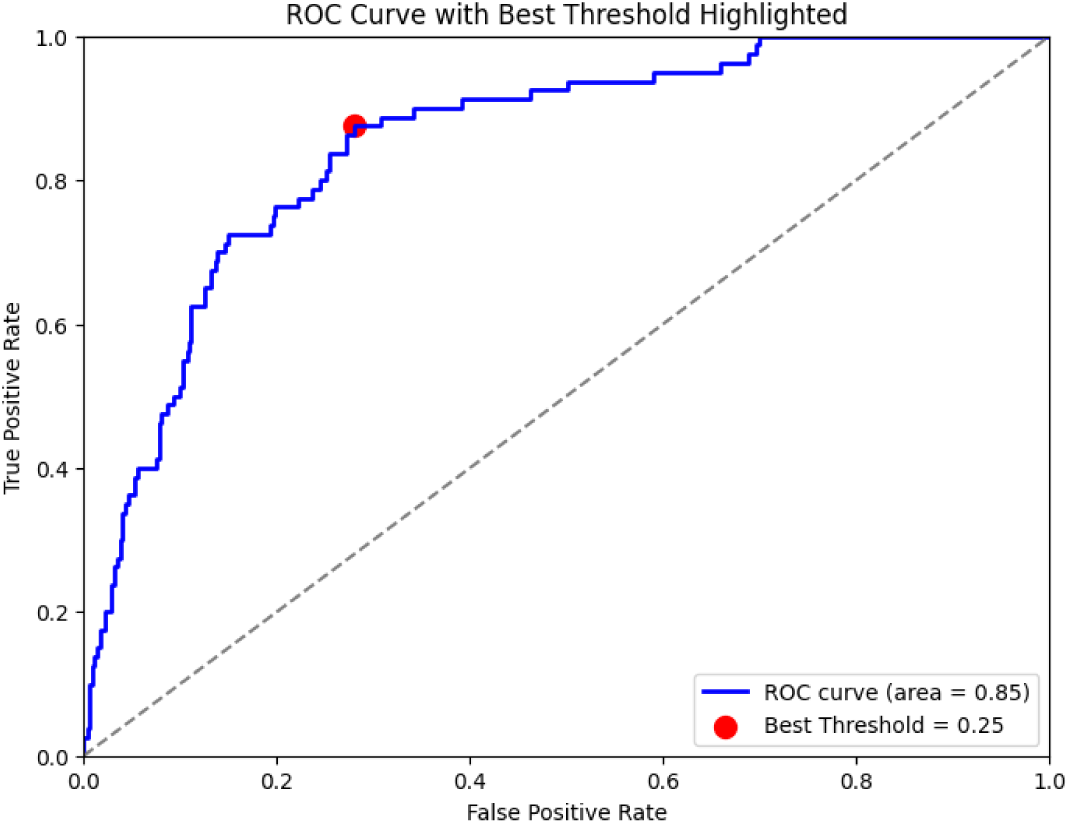
Receiver Operating Characteristic (ROC) curve based on the optimal risk threshold for predicting MACE in a FH population. The ROC curve depicts the performance of the predictive model using the selected optimal risk threshold for MACE. The curve plots sensitivity (true positive rate) against 1-specificity (false positive rate) at various threshold settings. The Area Under the Curve (AUC) reflects the model’s discriminative ability, with the optimal threshold chosen to balance sensitivity and specificity. This figure highlights the model’s accuracy in identifying patients at risk of MACE within the FH population.

Figure 5 shows the distribution of the FH population and the presence or absence of MACE based on the defined optimal risk threshold. Of the total samples, 38 of the samples exceeded this threshold, 61.4% of men and 22.7% of women with MACE were above this threshold.

**Fig. 5.**
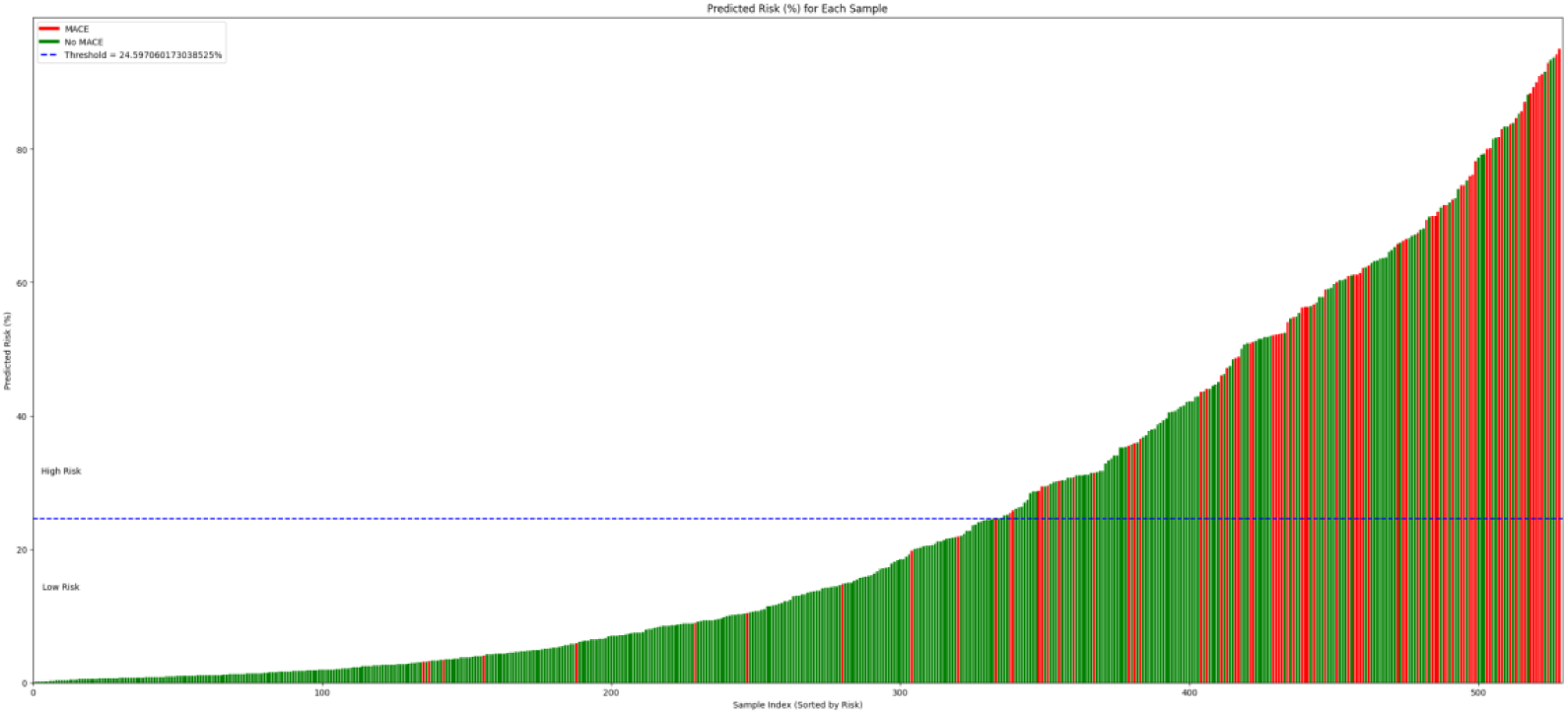
Distribution of patients based on the cardiovascular risk threshold. Each bar represents the risk of a patient and is ordered from lowest risk (left) to highest risk. The blue dashed line corresponds to the defined threshold of 0.25. Everything above it is classified as “High Risk,” while everything below is “Low Risk”. The red bars indicate MACE, and the green bars indicate non-MACE. Red bars below the threshold are false negatives, and green bars above the threshold are false positives.

### Analysis of the contribution of different variables in the AI/ML model, stratified by sex, using SHAP methodology

Figures 6 and 7 display the contribution of various variables in the model created using the SHAP methodology. In women, age, current GGT, the presence of subclinical disease, waist circumference, and Apoprotein B were the most influential factors in the model. In men, age, age of statin initiation, current HbA1c, LDL-cholesterol, and combination therapy had the greatest impact. The importance of each variable in the model differs significantly by sex. The results highlight sex-specific differences in the importance of key variables, suggesting the need for tailored predictive models that account for these variations.

**Fig. 6.**
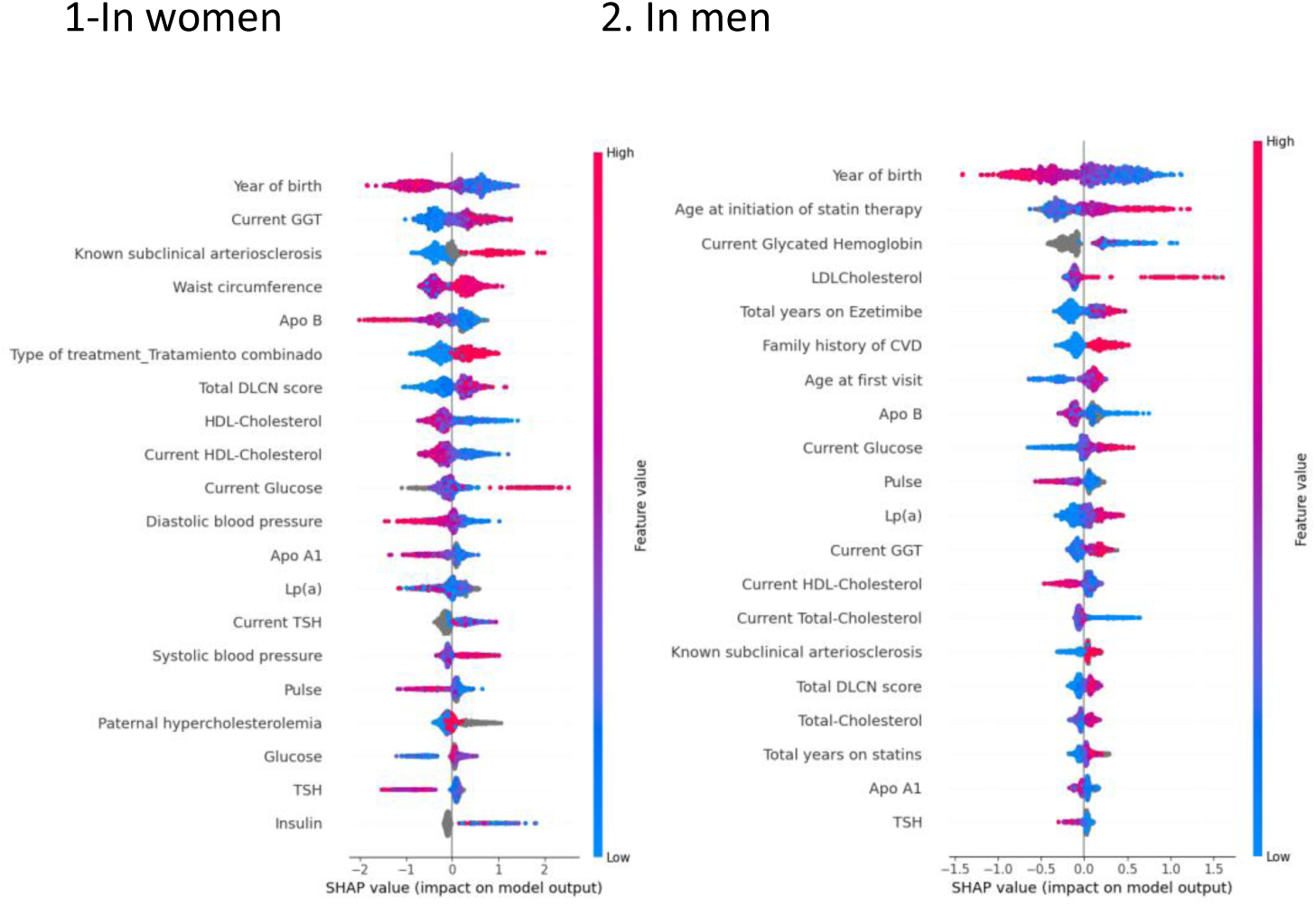
SHAP plot shows features that contribute to pushing the output from the base value of MACE by sex. The colored plot should be treated as a version of a violin plot where vertical height reflects number of observations with a given SHAP value, color of this vertical bar reflects the value of this specific variable for a given observation and horizontal distance from null reflects influence of this variable on the classification (further from the null means that this variable was more impactful on the estimated risk of this observation), positive SHAP value means predictor variable contributed to increased risk of this observation, negative SHAP value means predictor variable contributed to lowering the risk of a given observation.

**Fig 7.**
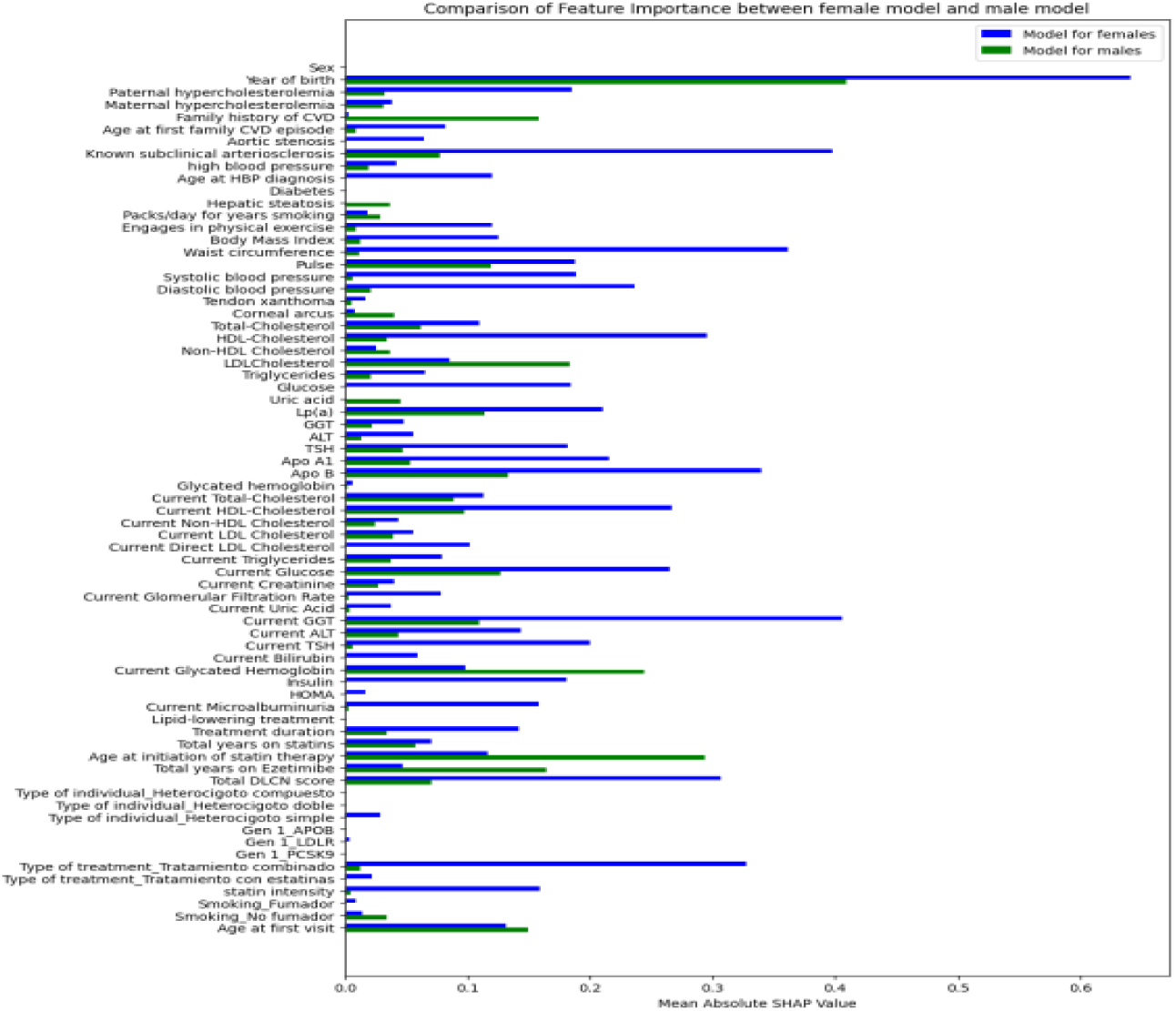
SHapley Additive exPlanations (SHAP) summary plot showing the contribution of individual variables to the prediction of MACE in FH patients. The SHAP plot illustrates the relative importance and impact of each variable on the model’s prediction of MACE, with separate analyses for male and female patients. Each bar represents the SHAP value for a given feature, where a higher SHAP value indicates a greater contribution to MACE risk prediction. Differences between sexes are emphasized, suggesting sex-specific predictors and interactions. This figure highlights the importance of personalized risk stratification by sex in FH populations.

## Discussion

To our knowledge, this work is the first study to evaluate an AI-ML algorithm applicable to the prognostic stratification of FH population from a sex-gender perspective.

The AI-ML algorithm developed for MACE risk in the FH population demonstrates strong predictive power that is superior to those previously obtained with inferencial multivariable models in the Spanish population (AI-ML: AUC 0.88 (95% CI 0.85-0.90); SAFEHEART-RE score: C-index 0.55 (95% CI 0.51-0.59) (9) and SIDIAP-HF score C-index:

0.71 (95% CI 0.68-0.75), (10). Indeed, it is superior to the metrics in the Canadian population (Montreal FH-score: AUC of 0.79 (95% CI 0.766-0.832) (8). The recall value in our model was moderately high (0.87 for the presence of MACE and 0.65 for the absence of MACE) but superior to the qualitative risk scales commonly used in clinical practice. In our study, 87.5% of the population with MACE, predominantly men exceeded the threshold detected for very high cardiovascular risk. In a recent analysis by, the SCORE Chart showed low sensitivity in Mediterranean patients with dyslipidemia. The scale classified 62.8% of the patients who experienced a cardiovascular event and 46.6% of those who died as low risk (26).

Understanding the heterogeneity in risk estimation and the role of emerging biomarkers and imaging techniques is crucial for optimizing cardiovascular risk prediction and guiding personalized treatment strategies in individuals with hypercholesterolemia. A combined approach using inferential statistics and AI techniques, incorporating data from different sources, is likely a good option at this time. In a newly released paper by Zinzuwadia et al, a machine learning approach enhanced the accuracy of the AHA-PREVENT model when applied to a local population while still preserving the risk associations identified by the original model. This strategy may help to reclassify patients into low or high cardiovascular risk categories (27).

Random Survival Forest (RSF) model is the most frequently utilised model for Survival Outcome in CVD Prediction. RSF is effective at handling complex interactions, has built-in variable importance measures, and is robust to overfitting. Despite RSL models often benefiting from large datasets, they can still be effectively applied to smaller health-related datasets as long as the right balance between data quantity and quality is ensured and interpretability is prioritised (28). Deep Learning models can also be useful in cardiovascular risk stratification, but the difficulty in explaining the model may lead to trust issues among professionals and patients (29). In a recent observational study conducted on a population in Bangladesh aged over 15 years, the best results in predicting cardiovascular events were achieved using the RSF, with an AUC of 0.98, compared to other ML-AI models (30). Other studies that have employed AI-ML techniques in the field of vascular risk stratification have also demonstrated improvements, particularly with the RSF, in diagnostic capability compared inferencial statistical methods, with AUCs ranging from 73 to 98 (31–36).

When the algorithm is evaluated in the subgroup divided by sex, we found different behavior in women and men. A recent study conducted in the SAFEHEART registry using inferential statistics has shown that the risk of ASCVD is markedly lower in females than in males with FH (37). In our IA developed model, it is observed that, generally, female sex is protective against the occurrence of cardiovascular events in the FH population. However,this protective effect disappeared, especially in younger women, when considering associations with other clinical or analytical characteristics.

Age, family history of hypercholesterolemia, aortic stenosis, the presence of subclinical disease, or waist circumference had a greater weight in the model for women, while family history of cardiovascular disease, hepatic steatosis, and the presence of corneal arcus had a greater weight in men. In the resulting model, an association is observed between the presence of MACE and LDL cholesterol, as well as the intensity of lipid-lowering treatment and the age at its initiation. However these variables have a greater impact in men than in women. In women, ApoB levels carry more weight in the model than LDL cholesterol levels. In a recent systematic review on the application of AI in cardiovascular risk stratification, limited evidence was found regarding sex differences (38). Of the 31 studies that included gender in their prediction models, only six studies performed gender-stratified predictions (39–44). None of them were conducted in a FH population. One of these studies did not observe differences by sex and race in the discriminative power of cardiovascular disease risk prediction between studies using neural networks and those conducted with pooled cohort equations when the same data were used for the analysis (42). High-dimensional features, including diverse sources such as clinical and laboratory data, genetics, social determinants, and imaging tests and/or longitudinal risk factors, evaluating variability between visits in laboratory values and vital signs, should be considered to fully explore the benefits of neural network survival models for cardiovascular risk prediction.

It is important that ML models provide intuitive explanations that enable patients to understand their risk predictions, thereby assisting clinicians and patients in better comprehending the decision-making process for assessing disease severity and maximizing opportunities for early intervention and personalized risk prediction models. Most prior studies have focused on the performance of ML models or the importance of features, with limited attention to fully understanding and explaining predictions using interpretable methods, such as SHAP (45), as in our study (46–47). Deep learning and neural network fields with larger datasets, related to cardiovascular disease, can be explored as future work, along with the integration of the trained machine learning models into proper explainable AI interface systems, such that the predictive results obtained from the machine learning models have a proper explainability, transparency, and are trusted bycitizens, patients, and clinicians (48).

The importance of this study lies in its demonstration that the use of AI-ML techniques can enhance prognostic stratification in FH populations compared to standard clinical practice. It applies a sex and gender perspective to avoid potential biases and includes explainability as a fundamental component. It is noteworthy that the data source is a national registry that includes a representative sample of patients from across the entire country with a significant number of participants. For the development of the model, different data from family history, clinical, analytical, genetic, imaging data and duration and intensity of lipid-lowering treatment were included.

This study has certain limitations. First, it is a cross-sectional study that provides a snapshot of information at a specific point in time, where the output is a prevalent variable rather than an incident one. To confirm the generalizability of these findings, future work will include validation in other claims and clinical data sets, ideally in a prospective study and clinical trials. Secondly, it includes an imbalanced population with lower representation of individuals with MACE, particularly women. On the other hand, there is likely to be an issue of collinearity among the different included variables.The algorithmic methods used have minimized these effects. Before potential implementation in clinical practice, it requires external validation in other populations, particularly in FH populations with phenotypic diagnosis. Future investigations could benefit from larger sample sizes to improve the robustness of our findings, ideally using large registries such as the European FH Patient Network (49) or CASCADE FH Registry of Family Heart Foundation (50), with special interest in pediatric and adolescent population, before widespread clinical implementation.The perspective of sex and ethnicity is crucial to avoid future biases and ensure an ethical approach to AI. In the future, integrating validated AI-ML algorithms into electronic health records (EHRs) in clinical practic, using criteria for robustness, transparency, ethics, and the inclusion of social determinants, as zip code, along with a sex-gender perspective, will enhance the management of diseases like FH, which represent a significant public health issue. Advancing towards an EHR with ’cognitive layers’ powered by AI could shift medicine from a reactive to a proactive approach. Ideally, screening and stratification algorithms should be embedded into healthcare systems’ electronic platforms to automatically detect citizens at very high vascular risk (’Electronic Red Flags’) and prioritize preventive and health promotion strategies. Moreover, this approach opens the door to new forms of epidemiological surveillance for non-communicable diseases (’Digital Epidemiology’), potentially generating ’high vascular risk maps’ to better target public health strategies. If we aim to avoid perpetuating the current sex biases in cardiovascular risk stratification in the new era of Medicine 4.0, we must prioritize the differences between men and women from the outset in the development of AI algorithms.

## Conclusion

AI-ML algorithms are promising tools for enhancing vascular risk stratification in patients with FH, revealing critical sex-based differences. Further validation in larger, more diverse populations, including prospective clinical trials, is the next step before widespread clinical implementation.

## Data Availability

The data included in this study were obtained from the National Registry of the Spanish Atherosclerosis Society.

http://www.rihad.es

## Acknowledgments

We would like to extend our gratitude to Dr. Adrià Pla for his valuable assistance in reviewing the manuscript for grammatical accuracy.

We would like to acknowledge the use of ChatGPT 4.0 as a writing assistant in the development of this manuscript, providing support in language refinement and grammar correction. The authors, however, take full responsibility for the content and scientific integrity of the work.

## Non-standard Abbreviations and Acronyms

AI: Artificial Intelligence
ASCVD: Atherosclerotic Cardiovascular Disease
AUC: Area Under the Curve
CAD: Coronary Artery Disease
CRISP-DM: Cross-Industry Standard Process for Data Mining
DLCN: Dutch Lipid Clinics Network
FH: Familial Hypercholesterolemia
HGBCT: Histogram-based Gradient Boosting Classification Tree
*K-FOLD*: *K-FOLD* cross-validation method
MACE: Major Adverse Cardiovascular Events
ML: Machine Learning
RIHAD: National Registry of Dyslipemias of the Spanish Society of Arteriosclerosis
SCORE: COronary Risk Evaluation Score
SEA: Spanish Society of Arteriosclerosis
SHAP: SHapley Additive exPlanations
ROC: Receiver Operating Characteristic
RSF: Random Survival Forest
XAI: Explainable Artificial Intelligence

## References

6 SCORE2-OP working group and ESC Cardiovascular risk collaboration. SCORE2-OP risk prediction algorithms: estimating incident cardiovascular event risk in older persons in four geographical risk regions. Eur Heart J. 2021 Jul 1;42(25):2455–2467. doi: 10.1093/eurheartj/ehab312.

7 Paquette M, Dufour R, Baass A. The Montreal-FH-SCORE: A new score to predict cardiovascular events in familial hypercholesterolemia. J Clin Lipidol. 2017 Jan-Feb;11(1):80-86. doi: 10.1016/j.jacl.2016.10.004.

8 Paquette M, Brisson D, Dufour R, Khoury É, Gaudet D, Baass A. Cardiovascular disease in familial hypercholesterolemia: Validation and refinement of the Montreal- FH-SCORE. J Clin Lipidol. 2017 Sep-Oct;11(5):1161–1167.e3. doi: 10.1016/j.jacl.2017.07.008.

9. Pérez de Isla L, Alonso R, Mata N, Fernández-Pérez C, Muñiz O, Díaz-Díaz JL, Saltijeral A, Fuentes-Jiménez F, de Andrés R, Zambón D et al. Predicting Cardiovascular Events in Familial Hypercholesterolemia: The SAFEHEART Registry (Spanish Familial Hypercholesterolemia Cohort Study). Circulation. 2017 May 30;135(22):2133–2144. doi: 10.1161/CIRCULATIONAHA.116.024541. Epub 2017 Mar 8. PMID: 28275165.

10 Ramos R, Masana L, Comas-Cufí M, García-Gil M, Martí-Lluch R, Ponjoan A, Plana N, Alves-Cabratosa L, Marrugat J, Elosua R, et al. Derivation and validation of SIDIAP-FHP score: A new risk model predicting cardiovascular disease in familial hypercholesterolemia phenotype. Atherosclerosis. 2020 Jan;292:42–51. doi: 10.1016/j.atherosclerosis.2019.10.016. Epub 2019 Oct 30. PMID: 31759248.

11 Feldman RD. Sex-Specific Determinants of Coronary Artery Disease and Atherosclerotic Risk Factors: Estrogen and Beyond. Can J Cardiol. 2020 May;36(5):706–711. doi: 10.1016/j.cjca.2020.03.002. Epub 2020 Mar 6. PMID: 32389343.

12 Klevmoen M, Mulder JWCM, Roeters van Lennep JE, Holven KB. Sex Differences in Familial Hypercholesterolemia. Curr Atheroscler Rep. 2023 Nov;25(11):861–868. doi: 10.1007/s11883-023-01155-6. Epub 2023 Oct 10. PMID: 37815650; PMCID: PMC10618303.

13 Jiménez A, Viñals C, Marco-Benedí V, González P, Domenech M, Suárez-Tembra M, Pinto X, Ortega E. Sex Disparities in FamilialHypercholesterolemia. J Am Coll Cardiol. 2023 Jan 17;81(2):203–205. doi: 10.1016/j.jacc.2022.10.023.

14 Zamora A, Ramos R, Comas-Cufi M, García-Gil M, Martí-Lluch R, Plana N, Alves-Cabratosa L, Ponjoan A, Rodríguez-Borjabad C, Ibarretxe D, et al. Women with familial hypercholesterolemia phenotype are undertreated and poorly controlled compared to men. Sci Rep. 2023 Jan 27;13(1):1492. doi: 10.1038/s41598-023-27963-z. PMID: 36707646; PMCID: PMC9883524.

15 Hossain S, Hasan MK, Faruk MO, Aktar N, Hossain R, Hossain K. Machine learning approach for predicting cardiovascular disease in Bangladesh: evidence from a cross-sectional study in 2023. BMC Cardiovasc Disord. 2024 Apr 18;24(1):214. doi: 10.1186/s12872-024-03883-2.

16 Luo RF, Wang JH, Hu LJ, Fu QA, Zhang SY, Jiang L. Applications of machine learning in familial hypercholesterolemia. Front Cardiovasc Med. 2023 Sep 26;10:1237258. doi: 10.3389/fcvm.2023.1237258. PMID: 37823179; PMCID: PMC10562581.

17 Teshale AB, Htun HL, Vered M, Owen AJ, Freak-Poli R. A Systematic Review of Artificial Intelligence Models for Time-to-Event Outcome Applied in Cardiovascular Disease Risk Prediction. J Med Syst. 2024 Jul 19;48(1):68. doi: 10.1007/s10916-024-02087-7. PMID: 39028429; PMCID: PMC11271333.

18 Pérez-Calahorra S, Sánchez-Hernández RM, Plana N, Valdivielso P, Civeira F. National Dyslipidemia Registry of the Spanish Arteriosclerosis Society: Current status. Clin InvestigArterioscler. 2017 Nov-Dec;29(6):248–253. English, Spanish. doi: 10.1016/j.arteri.2017.09.001. Epub 2017 Nov 2. PMID: 29102473.

19 Python Software Foundation. (n.d.). Python. Accesed October 5, 2024, from https://www.python.org/

20 Schröer, Christoph, Felix Kruse, and Jorge Marx Gómez. A systematic literature review on applying CRISP-DM process model. Procedia Computer Science, 2021: 181: 526–534. DOI: 10.1016/j.procs.2021.01.199

21 Nhat-Duc, Hoang; Van-Duc, Tran. Comparison of histogram-based gradient boosting classification machine, random Forest, and deep convolutional neural network for pavement raveling severity classification. Automation in Construction, 2023, vol. 148, p. 104767.

22 Dutschmann TM, Kinzel L, Ter Laak A, Baumann K. Large-scale evaluation of k-fold cross-validation ensembles for uncertainty estimation. J Cheminform. 2023 Apr 28;15(1):49. doi: 10.1186/s13321-023-00709-9.

23 Lundberg, S. A unified approach to interpreting model predictions. arXiv preprint arXiv:2017; 1705.07874.doi.org/10.48550/arXiv.1705.07874

24 Slack, Dylan, Sophie Hilgard, Emily Jia, Sameer Singh, and Himabindu Lakkaraju. “Fooling LIME and SHAP: Adversarial Attacks on Post Hoc Explanation Methods.” Proceedings of the AAAI/ACM Conference on Artificial Intelligence, Ethics, and Society (2020): 180–186. 10.48550/arXiv.1911.02508

25 Smits, N. A note on Youden’s *J*and its cost ratio. BMC Med Res Methodol, 2010; 1:89. 10.1186/1471-2288-10-89

26 Bertomeu-González V, Soriano Maldonado C, Bleda-Cano J, Carrascosa-Gonzalvo S, Navarro-Perez J, López-Pineda A, Carratalá-Munuera C, Gil Guillén VF, Quesada JA, Brotons C, et al. Predictive validity of the risk SCORE model in a Mediterranean population with dyslipidemia. Atherosclerosis. 2019 Nov;290:80–86. doi: 10.1016/j.atherosclerosis.2019.09.007. Epub 2019 Sep 23. PMID: 31593904.

27 Zinzuwadia AN, Mineeva O, Li C, Farukhi Z, Giulianini F, Cade B, Chen L, Karlson E, Paynter N, Mora S, Demler O. Tailoring Risk Prediction Models to Local Populations. JAMA Cardiol. 2024 Sep18:e242912. doi: 10.1001/jamacardio.2024.2912.

28 Ali, Sajid; Abuhmed, Tamer; El-Sappagh, Shaker; Muhammad, Khan; Alonso-Moral, Jose M.; Confalonieri, Roberto; Guidotti, Riccardo; Del Ser, Javier; Díaz-Rodríguez, Natalia; Herrera Triguero, Francisco. Explainable Artificial Intelligence (XAI): What we know and what is left to attain Trustworthy Artificial Intelligence. InformationFusion; 2023; 99; 101805.10.1016/j.inffus.2023.101805

29 Huang Y, Li J, Li M, Aparasu RR. Application of machine learning in predicting survival outcomes involving real-world data: a scoping review. BMC Med Res Methodol. 2023 Nov 13;23(1):268. doi: 10.1186/s12874-023-02078-1.

30 Hossain S, Hasan MK, Faruk MO, Aktar N, Hossain R, Hossain K. Machine learning approach for predicting cardiovascular disease in Bangladesh: evidence from a cross-sectional study in 2023. BMC Cardiovasc Disord. 2024 Apr 18;24(1):214. doi: 10.1186/s12872-024-03883-2.

31 Md. Imam Hossain, Mehadi Hasan Maruf, Md. Ashikur Rahman Khan, Farida Siddiqi Prity, Sharmin Fatema, Md. Sabbir Ejaz, Ahnaf Sad Khan. Heart disease prediction using distinct artificial intelligence techniques: performance analysis and comparison. Iran J Comput Sci. 2023; 6, 397–417 (2023). 10.1007/s42044-023-00148-7

32 Ali MM, Paul BK, Ahmed K, Bui FM, Quinn JMW, Moni MA. Heart disease prediction using supervised machine learning algorithms: Performance analysis and comparison. Comput Biol Med. 2021 Sep;136:104672. doi: 10.1016/j.compbiomed.2021.104672.

33 Kumar, N. K., Sindhu, G. S., Prashanthi, D. K., &Sulthana, A. S. Analysisan prediction of cardio vascular disease using machine learning classifiers. In 2020 6th International Conference on Advanced Computing and Communication Systems (ICACCS) (pp. 15–21). IEEE. DOI: 10.1109/ICACCS48705.2020.9074183

34 FAHIM, Khairul Eahsun, et al. Detection of Cardiovascular Disease of Patients at an Early Stage Using Machine Learning Algorithms. En 2022 International Conference on Healthcare Engineering (ICHE). IEEE, 2022. p. 1–6. DOI: 10.1109/ICHE55634.2022.10179871

35 M. D. Amzad Hossen, Tahia Tazin, Sumiaya Khan, Evan Alam, Hossain Ahmed Sojib, Mohammad Monirujjaman Khan, Abdulmajeed Alsufyani..Supervised Machine Learning-Based Cardiovascular Disease Analysis and Prediction. Mathematical Problems in Engineering, Hindawi, 2021; vol. 2021, pages 1-10: DOI: 10.1155/2021/1792201

36 Nashif, S., Raihan, Md.R., Islam, Md.R. and Imam, M.H. (2018) Heart Disease Detection by Using Machine Learning Algorithms and a Real-Time Cardiovascular Health Monitoring System. World Journal of Engineering and Technology, 6, 854–873. doi: 10.4236/wjet.2018.64057

37 de Isla LP, Vallejo-Vaz AJ, Watts GF, Muñiz-Grijalvo O, Alonso R, Diaz-Diaz JL, Arroyo-Olivares R, Aguado R, Argueso R, Mauri M, et al. Long-term sex differences in atherosclerotic cardiovascular disease in individuals with heterozygous familial hypercholesterolaemia in Spain: a study using data from SAFEHEART, a nationwide, multicentre, prospective cohort study. Lancet Diabetes Endocrinol. 2024 Sep;12(9):643–652. doi: 10.1016/S2213-8587(24)00192-X.

38 Teshale AB, Htun HL, Vered M, Owen AJ, Freak-Poli R. A Systematic Review of Artificial Intelligence Models for Time-to-Event Outcome Applied in Cardiovascular Disease Risk Prediction. J Med Syst. 2024 Jul 19;48(1):68. doi: 10.1007/s10916-024-02087-7. PMID: 39028429; PMCID: PMC11271333.

39 Barbieri S, Mehta S, Wu B, Bharat C, Poppe K, Jorm L, Jackson R. Predicting cardiovascular risk from national administrative databases using a combined survival analysis and deep learning approach. Int J Epidemiol. 2022 Jun 13;51(3):931–944. doi: 10.1093/ije/dyab258.

40 Blanchard M, Feuilloy M, Sabil A, Gerves-Pinquie C, Gagnadoux F, Girault JM. A Deep Survival Learning Approach for Cardiovascular Risk Estimation in Patients With Sleep Apnea. IEEE Access. 2022;10:133468–78. DOI: 10.1109/ACCESS.2022.3231743

41 Chun M, Clarke R, Cairns BJ, Clifton D, Bennett D, Chen Y, Guo Y, Pei P, Lv J, Yu C, Yang L, Li L, Chen Z, Zhu T; China Kadoorie Biobank Collaborative Group. Stroke risk prediction using machine learning: a prospective cohort study of 0.5 million Chinese adults. J Am Med Inform Assoc. 2021 Jul 30;28(8):1719–1727. doi: 10.1093/jamia/ocab068.

42 Deng Y, Liu L, Jiang H, Peng Y, Wei Y, Zhou Z, Zhong Y, Zhao Y, Yang X, Yu J, Lu Z, Kho A, Ning H, Allen NB, Wilkins JT, Liu K, Lloyd-Jones DM, Zhao L. Comparison of State-of-the-Art Neural Network Survival Models with the Pooled Cohort Equations for Cardiovascular Disease Risk Prediction. BMC Med Res Methodol. 2023 Jan 24;23(1):22. doi: 10.1186/s12874-022-01829-w.

43 Qian X, Keerman M, Zhang X, Guo H, He J, Maimaitijiang R, Wang X, Ma J, Li Y, Ma R, Guo S. Study on the prediction model of atherosclerotic cardiovascular disease in the rural Xinjiang population based on survival analysis. BMC Public Health. 2023 Jun 1;23(1):1041. doi: 10.1186/s12889-023-15630-x.

44 Sung JM, Cho IJ, Sung D, Kim S, Kim HC, Chae MH, Kavousi M, Rueda-Ochoa OL, Ikram MA, Franco OH, Chang HJ. Development and verification of prediction models for preventing cardiovascular diseases. PLoS One. 2019 Sep 19;14(9):e0222809. doi: 10.1371/journal.pone.0222809.

45 Salah H, Srinivas S. Explainable machine learning framework for predicting long-term cardiovascular disease risk among adolescents. Sci Rep. 2022 Dec 19;12(1):21905. doi: 10.1038/s41598-022-25933-5.

46 Sharma, S.; Parmar, M. Heart diseases prediction using deep learning neural network model. Int. J. Innov. Technol. Explor. Eng. 2020, 9, 124–137. DOI: 10.35940/ijitee.C9009.019320

47 Miranda E, Adiarto S, Bhatti FM, Zakiyyah AY, Aryuni M, Bernando C. Understanding Arteriosclerotic Heart Disease Patients Using Electronic Health Records: A Machine Learning and Shapley Additive exPlanations Approach. Healthc Inform Res. 2023 Jul;29(3):228–238. doi: 10.4258/hir.2023.29.3.228.

48 Guleria, P.; Naga Srinivasu, P.; Ahmed, S.; Almusallam, N.; Alarfaj, F.K. XAI Framework for Cardiovascular Disease Prediction Using Classification Techniques. Electronics 2022, 11, 4086. 10.3390/electronics11244086

49 Bedlington N, Abifadel M, Beger B, Bourbon M, Bueno H, Ceska R, Cillíková K, Cimická Z, Daccord M, de Beaufort C, et al. The time is now: Achieving FH paediatric screening across Europe - The Prague Declaration. GMS Health Innov Technol. 2022 Sep 30;16:Doc04. doi: 10.3205/hta000136.

50 Representatives of the Global Familial Hypercholesterolemia Community; Wilemon KA, Patel J, Aguilar-Salinas C, Ahmed CD, Alkhnifsawi M, Almahmeed W, Alonso R, Al- Rasadi K, Badimon L, Bernal LM, et al. Reducing the Clinical and Public Health Burden of Familial Hypercholesterolemia: A Global Call to Action. JAMA Cardiol. 2020 Feb 1;5(2):217–229. doi: 10.1001/jamacardio.2019.5173. Erratum in: JAMA Cardiol. 2020 May 1;5(5):613. doi: 10.1001/jamacardio.2020.0344. PMID: 31895433.

